# Whole exome sequencing in multi-incident families identifies novel candidate genes for multiple sclerosis

**DOI:** 10.1101/2022.02.28.22271609

**Authors:** J. Horjus, T. Banda, M. Heerings, M. Hakobjan, W. De Witte, D.J. Heersema, A.J. Jansen, E.M.M. Strijbis, B.A. de Jong, A.E.J. Slettenaar, E.M.P.E. Zeinstra, E.L.J. Hoogervorst, B. Franke, W. Kruijer, P.J. Jongen, L. Visser, G. Poelmans

**Affiliations:** Department of Human Genetics, Radboud University Medical Center, Nijmegen, The Netherlands; Department of Neurology, University Medical Center Groningen, University of Groningen, Goningen, The Netherlands; Department of Neurology, Bravis Hospital, Bergen op Zoom, The Netherlands; Department of Neurology, Amsterdam UMC, location VUmc, Amsterdam, The Netherlands; Department of Neurology, Amsterdam UMC, Vrije Universiteit Amsterdam, MS Center Amsterdam, Amsterdam Neuroscience, Amsterdam, The Netherlands; Department of Neurology, Medisch Spectrum Twente, Enschede, The Netherlands; Department of Neurology, Isala Hospital, Meppel, The Netherlands; Department of Neurology, St Antoniusziekenhuis, Utrecht/Nieuwegein, the Netherlands; Donders Institute for Brain, Cognition and Behaviour, Nijmegen, The Netherlands; Department of Psychiatry, Radboud University Medical Center, Nijmegen, The Netherlands; Independent Life Science Consultant, Leusden, The Netherlands; MS4 Research Institute, Nijmegen, The Netherlands; Department of Community & Occupational Medicine, University of Groningen, University Medical Centre Groningen, Groningen, The Netherlands; Department of Neurology, St. Elisabeth-Tweesteden Hospital, Tilburg, The Netherlands; Department of Care Ethics, University of Humanistic Studies, Utrecht, The Netherlands

## Abstract

Multiple sclerosis (MS) is a degenerative disease of the central nervous system in which auto-immunity-induced demyelination occurs. MS is thought to be caused by a complex interplay of environmental and genetic risk factors. While most genetic studies have focused on identifying common genetic variants for MS through genome-wide association studies, the objective of the present study was to identify rare genetic variants contributing to MS susceptibility. We used whole exome sequencing (WES) followed by co-segregation analyses in nine multi-incident families with 2 to 4 affected individuals. WES was performed in 31 family members with and without MS. After applying a suite of selection criteria, co-segregation analyses for a number of rare variants selected from the WES results were performed, adding 24 family members. This approach resulted in 12 exonic rare variants that showed acceptable co-segregation with MS within the nine families, implicating the genes *MBP, PLK1, MECP2, MTMR7, TOX3, CPT1A, SORCS1, TRIM66, ITPR3, TTC28, CACNA1F*, and *PRAM1*. Of these, three genes (*MBP, MECP2*, and *CPT1A*) have been previously reported as carrying MS-related rare variants. Six additional genes (*MTMR7, TOX3, SORCS1, ITPR3, TTC28*, and *PRAM1*) have also been implicated in MS through common genetic variants. The proteins encoded by all twelve genes containing rare variants interact in a molecular framework that points to biological processes involved in (de-/re-)myelination and auto-immunity. Our approach provides clues to possible molecular mechanisms underlying MS that should be further studied in cellular and/or animal models.

## Introduction

Multiple Sclerosis (MS) is one of the most common neurological diseases, affecting over two million people globally and showing an increasing incidence ^1^. Auto-immunity pathways play an important role in the pathogenesis of MS ^2^. The focal areas of inflammation associated with MS are often characterized by reduced integrity of the blood-brain barrier (BBB), leading to infiltration of several types of immune cells, including myelin-specific autoreactive T-cells and macrophages ^3,4^. This auto-immune response results in the activation of microglia, which in turn promotes further neuroinflammation and demyelination through the release of proinflammatory cytokines ^5^. Additionally, proinflammatory mediators released by activated T-cells and microglia cause damage to oligodendrocytes (ODCs) that produce and maintain the myelin sheaths around axons in the central nervous system (CNS) ^6^. The MS-associated demyelinating events in the CNS and the inability to remyelinate the resulting lesions eventually lead to axonal and neuronal degeneration ^7^. Depending on the extent and location of the MS lesions, a variety of symptoms can occur, such as motor, sensory, cognitive, and visual symptoms and, eventually, severe overall disability.

In case of remyelination, which is a normal process in healthy individuals, the underlying axons remain intact and neurodegeneration is prevented. Remyelination is achieved through the differentiation of progenitor cells into mature ODCs ^8^. Microglia and macrophages, which are also able to release anti-inflammatory cytokines in addition to the abovementioned proinflammatory cytokines ^9^, have been found to stimulate ODC differentiation from progenitor cells, therefore contributing to remyelination ^10^. Remyelination can occur in the remission phases in the relapsing-remitting form of MS (RRMS), the (initial) subtype found in approximately 90% of MS cases ^11^. In RRMS, relapses characterized by (various) neurological symptoms due to inflammatory and active demyelinating lesions alternate with periods in which these symptoms are partially or completely absent. In 15-80% of MS cases, the disease course becomes progressive within 10 to 15 years, which is called secondary progressive MS (SPMS) ^12,13^. The small group of patients with primary progressive MS (PPMS) experiences uninterrupted progression from disease onset.

Progressive forms of MS are pathologically characterized by brain atrophy, which can be linked to axonal and neuronal degeneration due to the lack of remyelination ^14^.

MS is thought to be caused by a complex interplay of multiple environmental and genetic risk factors that contribute to disease onset and progression ^15^. Environmental risk factors for MS include Epstein-Barr virus infection ^16^, vitamin D deficiency, and cigarette smoking ^17^. Females are affected up to three times more often than males, and this is assumed to be largely due to environmental factors ^18^. Furthermore, first-degree relatives of patients have a 20–40-fold risk of developing MS compared to the general population, and this risk is 300-fold for monozygotic twins ^19^. The genetic contribution to MS susceptibility seems to be in part polygenic, i.e. a large number of common variants within multiple genes each have a small contribution to overall disease risk ^20^. Large-scale genome-wide association studies (GWASs) have shown that the common genetic risk for MS is for the largest part due to variance in the Major Histocompatibility Complex (MHC) gene cluster, including multiple human leucocyte antigen (*HLA*) genes ^21^. These genes encode proteins with important immunological functions, which is in line with the characterization of MS as an auto-immune disease. A total of 20-30% of MS heritability can be explained by common variants within *HLA* genes ^22,23^. The most recent and largest meta-analysis of GWASs of MS analysed 8,278,136 common genetics variants (i.e., single nucleotide polymorphisms, SNPs) in 47,429 MS cases and 68,374 controls. This meta-GWAS identified 233 SNPs that showed genome-wide significant association with MS (P < 5.00E-08), including 200 autosomal SNPs in genes outside the MHC cluster and one SNP on the X-chromosome ^23^.

In addition to common genetic variants, rare variants have been shown to contribute to MS risk. Next-generation sequencing methods, especially whole exome or whole genome sequencing (WES or WGS), are considered to be the elementary technology for the discovery of rare variants involved in diseases, but exome chips are also frequently used. In this respect, a large exome chip study of 120,991 coding low-frequency variants with minor allele frequencies (MAFs) between 0.01% and 5% compared 32,367 MS cases and 36,012 controls ^24^.

The results of this study showed that up to 5% of MS heritability is explained by low-frequency variants. Some of these variants were found to impact genes that have not yet been observed in GWASs and that encode proteins involved in diverse immunity-related processes, such as regulatory T cell homeostasis, IFNγ biology, and NFkB signaling ^24^.

WES and WGS studies of rare variants in MS generally use family designs. When multiple family members are affected within a family, rare variants contributing to disease risk can be considered to be present at higher frequencies compared to the general population ^25^. In addition, accumulation of rare susceptibility variants is observed in multi-incident MS families compared to single-case families ^26^. This makes multi-incident MS families of interest for identifying and studying rare variants involved in the disease. In the past 10 years, a number of studies have reported rare, putatively pathogenic genetic variants in MS patients from multi-incident families through WES ^22,27-32^. Most of these family studies, however, only involved a small number of families, analyses were sometimes targeted exclusively at known MS loci, and the vast majority of the identified variants were not studied in replication studies or did not replicate ^27,29,33-36^. Therefore, more robust WES/WGS studies and integration across studies of common and rare genetic variant findings are still needed to better understand the rare genetic component of MS and provide additional insight into the molecular pathways underlying the disease.

In the present study, WES and co-segregation analyses were applied in nine Dutch MS families with 2 to 4 affected family members, in order to identify rare, exonic variants co-segregating with the disease in these families. Our findings were integrated with the results from large rare and common variant studies of MS. Subsequently, based on the proteins encoded by the genes in which we identified rare variants, we constructed a molecular framework that provides further insights into the mechanisms underlying MS as a first step towards validation studies in cellular and/or animal models.

## Methods

### Participants

Multi-incident MS families of Dutch descent were recruited through an online advertisement on the website of the Dutch National MS Foundation (that also funded this study). In order to participate in this study, at least two people with MS needed to be present in a family, preferably across two generations. Participants had to be minimally 20 years of age and have a diagnosis of MS - made by a neurologist - according to the McDonald criteria for at least one year ^37^. After online application, families were contacted by phone to plan a home visit. At this home visit, blood or saliva for DNA isolation was collected from the MS cases and as many unaffected family members as possible. In addition, informed consent was obtained and a questionnaire was administered to collect information about disease type and duration, MS-related treatments and more general information such as (previous) infectious and other diseases in both MS cases and unaffected family members. Lastly, a pedigree of the family was drawn to obtain more information about the disease penetrance. A total of nine multi-incident MS families were included. The ‘Medische Ethische Toetsingscommissie (METC) Brabant’ – i.e., the relevant Dutch Medical Ethical Committee – approved this study, and written informed consent was provided by each participant (approved research study number: NL62481.028.17 / P1738).

### Whole exome sequencing and analysis

DNA was available for a total of 55 members across the nine families, of which 26 were diagnosed with MS and 29 were unaffected family members. Genomic DNA was isolated from either peripheral venous blood or saliva according to standard procedures of the department of Human Genetics of the Radboud University Medical Center, Nijmegen, The Netherlands. Whole exome sequencing was performed by BGI (Copenhagen, Denmark) on at least two MS cases per family and one unaffected family member. Exome capture was performed by using the SureSelect^XT^ Human All Exon V5 enrichment kit (Agilent Technologies, Santa Clara, CA, USA).

Raw image files were generated on HiSeq 4000 platform (Illumina, San Diego, CA, USA), making use of the 2x 100 bp paired-end module with a minimal median target coverage of 50X and an average Phred-like consensus quality of 30 for at least 80% of called bases. This implies that 80% of called bases have an accuracy of 99.9%. Base calling was implemented through the default parameters of the Illumina software. Sequencing reads were then aligned against the human reference genome GRCh37/hg19 using the Burrows-Wheeler Aligner (BWA). Variants were subsequently called by the Genome Analysis Toolkit (GATK) HaplotypeCaller. Annotation of variants was performed according to in-house pipelines of the department of Human Genetics. Data on 31 sequenced exomes were available from this procedure.

### Selection of candidate single nucleotide variants

For each family, rare single nucleotide variants (SNVs) in the exome were selected that were present in all sequenced MS cases but absent in the sequenced unaffected family member(s), making use of the in-house tool HitPanda. In order to identify rare variants most likely involved in the pathogenesis of MS in each family, additional selection procedures were applied.

Firstly, SNVs occurring in at least two families were selected. Of these SNVs, only those identified in ≤1% of the general population were selected. Both non-synonymous substitutions – ‘missense mutations’ – and other (putative) loss-of-function variants were included. In addition, variants with a Combined Annotation Dependent Depletion (CADD) score below 20 were omitted. The CADD score provides a measurement of the deleteriousness of a variant; a score of 20 implies that the variant belongs to the 1% most deleterious substitutions in the human genome ^38^.

As a second selection strategy, a selection was made among variants within single families. Only rare SNVs identified in ≤0.05% of the general population were included, using the same criteria as described above. In addition to missense variants, all putative loss-of-function variants fulfilling these criteria were included. Among the selected missense variants, a further selection was made based on prior evidence.

Variants within genes for which literature evidence of a clear link with MS could be found were selected. Additionally, variants were selected based on evidence of association in the abovementioned large exome chip ^24^ and GWAS ^23^ analyses from the International Multiple Sclerosis Genetics Consortium (IMSGC). Using the summary statistics of the GWAS, downloaded from the website of the IMSGC (https://imsgc.net/), we performed a gene-wide association analysis using the Multimarker Analysis of GenoMic Annotation (MAGMA) software ^39^. From this, a weighted P-value for the combination of all SNPs within a gene was calculated (see **Supplementary Table 1** for all genes with uncorrected P-values <0.05). We required variants from the exome study and genes from the gene-wide GWAS to show association at uncorrected P<0.05 for selection of a WES-derived variant for further analysis.

### Validation of SNVs

Validation was performed by Sanger sequencing of the region of interest for each variant in the respective family. The region of interest was sequenced in all family members with DNA available, including the 24 family members in which WES had not been performed. Primer pairs spanning the variant of interest were designed according to in-house pipelines, making use of the University of California Santa Cruz (UCSC) Genome Browser and Primer3Plus online software. Primer optimization was achieved by running a gradient polymerase chain reaction (PCR) on test DNA to determine the optimal annealing temperature for each primer pair. Primer sequences and annealing temperatures can be found in **Supplementary Table 2**. The purified PCR product from the participating family members was then sequenced at the sequencing facility of the department of Human Genetics. Sequences were analyzed using Vector NTI software (Invitrogen, Carlsbad, CA, USA) and Alamut Visual v2.10 software (Interactive Biosoftware, Rouen, France).

Co-segregation analysis was then performed to assess to which extent MS cases in a given family carried the identified rare variant, while unaffected family members did not.

We designated those variants that were present in all MS cases and <33% of unaffected family members as showing ‘acceptable’ co-segregation.

### Construction of molecular framework

Subsequently, we constructed a molecular framework based on the proteins encoded by genes containing rare variants that met our abovementioned selection criteria and showed acceptable co-segregation. In short, we conducted an elaborate literature analysis of the (putative) function of these proteins, their interactions and evidence of these genes/proteins being implicated in MS. In addition, we placed some other proteins that have been implicated in MS and functionally link the proteins from our WES in the framework. Based on much larger data sets, we have previously applied a similar approach to build so-called ‘molecular landscapes’ for other neurological disorders, such as Parkinson’s disease ^40^ and amyotrophic lateral sclerosis ^41^.

## Results

Nine families with 2 to 4 MS cases were included in this study. The characteristics of the 55 participants (26 MS patients and 29 unaffected family members) are presented in **Table 1**. The mean age at DNA collection was 45.6 ± 15.2 years for MS cases and 51.2 ± 16.0 years for the unaffected family members. The average age at MS onset was 31.6 ± 8.5 years. The male to female ratio was 1:2.9 for MS cases and 1:1.2 for unaffected family members. All forms of MS were observed within and across the families (RRMS, n=13; SPMS, n=6, PPMS, n=2, diagnosis made by a neurologist), while for five cases, the MS form/disease course was not possible to determine.

**Table 1.** An overview of the clinical characteristics of the participants included in the whole exome sequencing (WES) study of MS (26 MS patients and 29 unaffected family members)/ F: female; M: male; NA: MS type not available; RRMS: relapsing-remitting MS; SPMS: secondary progressive MS; PPMS: primary progressive MS.

| Family | Participant | Gender | MS diagnosis | MS subtype | Age range at diagnosis (years) | Age range at DNA collection (years) | WES performed |
| --- | --- | --- | --- | --- | --- | --- | --- |
| 1 | II.1 | F | No | - | - | 61-65 | No |
| 1 | II.4 | F | No | - | - | 36-40 | Yes |
| 1 | II.5 | M | Yes | RRMS | 26-30 | 36-40 | Yes |
| 1 | II.11 | F | Yes | RRMS | 26-30 | 31-35 | No |
| 1 | II.13 | F | Yes | RRMS | 26-30 | 26-30 | Yes |
| 2 | II.1 | M | No | - | - | 81-85 | Yes |
| 2 | III.1 | M | No | - | - | 56-60 | No |
| 2 | III.2 | F | Yes | SPMS | 46-50 | 56-60 | Yes |
| 2 | III.10 | F | No | - | - | 51-55 | No |
| 2 | III.12 | F | Yes | SPMS | 16-20 | 41-45 | Yes |
| 2 | IV.1 | F | No | - | - | 26-30 | No |
| 2 | IV.2 | M | Yes | RRMS | 21-25 | 21-25 | No |
| 3 | III.1 | M | Yes | RRMS | 21-25 | 31-35 | Yes |
| 3 | II.1 | M | No | - | - | 56-60 | Yes |
| 3 | II.2 | F | Yes | RRMS | 31-35 | 56-60 | Yes |
| 3 | II.3 | M | No | - | - | 51-55 | No |
| 3 | III.3 | F | No | - | - | 31-35 | No |
| 4 | III.2 | M | Yes | SPMS | 51-55 | 71-75 | Yes |
| 4 | III.3 | F | No | - | - | 66-70 | Yes |
| 4 | III.6 | F | No | - | - | 61-65 | No |
| 4 | III.8 | F | Yes | NA | 21-25 | 56-60 | No |
| 4 | IV.1 | F | No | - | - | 31-35 | No |
| 4 | IV.2 | F | Yes | RRMS | 26-30 | 31-35 | Yes |
| 4 | IV.4 | F | Yes | RRMS | 31-35 | 31-35 | Yes |
| 4 | IV.5 | M | No | - | - | 46-50 | No |
| 4 | IV.6 | M | No | - | - | 26-30 | No |
| 4 | IV.7 | F | No | - | - | 21-25 | No |
| 5 | I.1 | M | Yes | SPMS | 26-30 | 66-70 | Yes |
| 5 | I.2 | F | No | - | - | 61-65 | No |
| 5 | II.1 | F | No | - | - | 36-40 | No |
| 5 | II.2 | M | Yes | RRMS | 36-40 | 36-40 | Yes |
| 5 | II.3 | F | Yes | RRMS | 26-30 | 36-40 | Yes |
| 5 | II.4 | M | No | - | - | 36-40 | Yes |
| 6 | I.1 | F | No | - | - | 61-65 | Yes |
| 6 | II.1 | F | Yes | RRMS | 26-30 | 31-35 | Yes |
| 6 | II.2 | F | No | - | - | 26-30 | No |
| 6 | II.3 | F | Yes | RRMS | 21-25 | 26-30 | Yes |

| Table 1 – continued. |  |  |  |  |  |  |  |
| --- | --- | --- | --- | --- | --- | --- | --- |
| Family | Participant | Gender | MS diagnosis | MS subtype | Age range at diagnosis (years) | Age range at DNA collection (years) | WES performed |
| 7 | II.2 | F | Yes | SPMS | 36-40 | 66-70 | Yes |
| 7 | II.5 | M | No | - | - | 61-65 | No |
| 7 | II.7 | F | Yes | SPMS | 31-35 | 61-65 | No |
| 7 | II.9 | F | No | - | - | 61-65 | No |
| 7 | III.1 | F | Yes | PPMS | 36-40 | 36-40 | Yes |
| 7 | III.2 | F | Yes | NA | 21-25 | 36-40 | Yes |
| 7 | III.3 | F | No | - | - | 36-40 | Yes |
| 7 | II.10 | M | No | - | - | 56-60 | No |
| 8 | I.1 | M | No | - |  | 66-70 | No |
| 8 | I.2 | F | No | - |  | 66-70 | No |
| 8 | II.1 | M | Yes | NA | 31-35 | 41-45 | Yes |
| 8 | II.2 | F | Yes | NA | 21-25 | 36-40 | Yes |
| 8 | II.3 | M | No | - |  | 36-40 | Yes |
| 8 | II.4 | F | Yes | RRMS | 31-35 | 36-40 | Yes |
| 9 | II.7 | F | Yes | NA | 41-45 | 76-80 | Yes |
| 9 | II.8 | F | Yes | PPMS | 51-55 | 71-75 | Yes |
| 9 | III.1 | M | No | - | - | 56-60 | No |
| 9 | III.2 | M | No | - | - | 51-55 | Yes |

Whole exome sequencing of 31 individuals from nine MS families resulted in 6.9 gigabases of mapped reads. Average target coverage was 80-fold per exome. An average of 122,969 variants was called per individual (range 115,583 – 130,153). Of these variants, 94.1% was annotated in dbSNP (v150).

Selection of the variants that were present in all sequenced MS cases of a family and absent in the unaffected sequenced family members resulted in an average total of 17,050 variants per family (range 7,636 – 23,881). Applying the described selection criteria, we obtained 15 missense variants and 3 deletions across all families, of which five missense variants were present in two out of nine families. In **Table 2**, the 18 variants are listed with frequencies, nucleotide and amino acid changes, and CADD scores. Rare missense variants in the genes *PLK1, PRAM1, CACNA1F, CPT1A*, and *TRIM66* were identified in two families. Among the 18 variants, missense variants were found in three genes that have been previously linked to MS: *MBP* ^42,43^, *MECP2* ^44,45^, and *CPT1A* ^46^. The missense variants in *PRAM1* and *USH2A* showed nominally significant association with MS in the exome chip study of the IMSGC ^24^. Furthermore, at least nominally significant gene-wide P-values were found in the data from the GWAS of the IMSGC for additional genes carrying rare variants in our study, i.e. *ZNF641, EXOC2, CCNI, ITPR3, SORCS1, PRAM1, MTMR7, TTC28*, and *TOX3* ^23^. An overview of all 18 variants meeting the selection criteria with accompanying P-values (where applicable) can be found in **Supplementary Table 3**.

**Table 2.** Rare genetic variants selected for validation. An overview of the variants that were selected according to our criteria for nine multi-incident MS families. All variants are non-synonymous substitutions, resulting in missense variants, apart from three loss-of-function variants constituting a deletion (DEL) of 7-15 nucleotides. Variants with a reference SNP identifier (rs ID) are known to NCBI’s dbSNP database (v150). For these variants, minor allele frequencies (MAFs) are mentioned. If a variant is novel, frequencies are indicated as NA (not available). Both nucleotide and amino acid changes are provided for each identified variant (where applicable), as well as the Combined Annotation Dependent Depletion score (CADD). The criteria for selection of each of the individual variants are shown in **Supplementary Table 3**.

| Family | Chromosome | Gene | Variant ID | MAF (%) | Nucleotide change | Amino acid change | CADD |
| --- | --- | --- | --- | --- | --- | --- | --- |
| 1 | chr6 | *EXOC2* | rs760365995 | ≤ 0.05 | G>A | p.Pro134Leu | 25.0 |
| 1 | chr16 | *PLK1* | rs35056440 | ≤ 1 | C>T | p.Leu261Phe | 22.9 |
| 1 | chr18 | *MBP* | 18:74696828 | NA | C>A | p.Gly151Val | 25.4 |
| 2 | chr12 | *ZNF641* | rs200502528 | ≤ 0.05 | G>A | p.His342Tyr | 27.7 |
| 3 | chr4 | *CCNI* | rs139547927 | ≤ 0.05 | T>C | p.Tyr334Cys | 29.1 |
| 3 | chr 6 | *ITPR3* | 6: 33648383 - 33648394 | NA | DEL (a) | - | 26.6 |
| 3 | chr10 | *SORCS1* | 10:108439392 | NA | G>A | p.Ser554Leu | 34.0 |
| 3 | chr16 | *PLK1* | rs35056440 | ≤ 1 | C>A | p.Leu261Phe | 22.9 |
| 3 | chr19 | *PRAM1* | rs138042924 | ≤ 1 | A>G | p.Ser387Pro | 22.0 |
| 3 | chrX | *CACNA1F* | X:49067910 | NA | T>A | p.Met1324Leu | 23.4 |
| 3 | chrX | *MECP2* | rs61751445 | ≤ 0.05 | G>A | p.Thr311Met | 26.1 |

| Table 2 – continued. |  |  |  |  |  |  |  |
| --- | --- | --- | --- | --- | --- | --- | --- |
| Family | Chromosome | Gene | Variant ID | MAF (%) | Nucleotide change | Amino acid change | CADD |
| 4 | chr1 | *USH2A* | rs200802261 | ≤ 0.05 | G>A | p.Pro2870Leu | 31.0 |
| 5 | chr11 | *CPT1A* | rs140958507 | ≤ 1 | C>T | p.Arg288Gln | 23.7 |
| 6 | chr8 | *MTMR7* | rs760803217 | ≤ 0.05 | G>A | p.Thr6Met | 22.5 |
| 6 | chr11 | *TRIM66* | rs138444298 | ≤ 1 | G>A | p.Arg1037Trp | 21.5 |
| 6 | chr16 | *CNGB1* | rs776392588 | ≤ 0.05 | DEL (b) | - | 51.0 |
| 7 | chrX | *CACNA1F* | X:49067910 | NA | T>A | p.Met1324Leu | 23.4 |
| 8 | chr22 | *TTC28* | 22:28501278 | NA | T>C | p.Tyr1099Cys | 32.0 |
| 9 | chr4 | *MANBA* | rs1283991082 | ≤ 0.05 | DEL (c) | - | 30.0 |
| 9 | chr11 | *TRIM66* | rs138444298 | ≤ 1 | G>A | p.Arg1037Trp | 21.5 |
| 9 | chr11 | *CPT1A* | rs140958507 | ≤ 1 | C>T | p.Arg288Gln | 23.7 |
| 9 | chr16 | *TOX3* | 16:52580565 | NA | T>C | p.Tyr24Cys | 23.8 |
| 9 | chr19 | *PRAM1* | rs138042924 | ≤ 1 | A>G | p.Ser387Pro | 22.0 |

Sanger sequencing was performed in all family members with available DNA in order to validate the WES results and to genotype the 24 family members in whom WES had not been performed. As shown in **Table 3** and **Supplementary Figure 1** - containing the pedigrees of the nine families with all 18 variants; as this figure may contain identifying information, it is only available upon request from the corresponding author - subsequent co-segregation analyses of each variant in the respective families indicated that the variants within five genes completely co-segregated with MS: *PLK1, MBP, SORCS1, MTMR7*, and *TOX3*. The identified variants in *PRAM1, CPT1A, TRIM66, CACNA1F, MECP2, TTC28*, and *ITPR3* were present in all MS cases in the respective families, but were also present in 20-33% of unaffected family members. The missense variants in *EXOC2* and *CCNI*, and the deletions in *CNGB1* and *MANBA*, were present in 50-66% of the unaffected individuals. Lastly, the missense variants in *ZNF641* and *USH2A* were not present in all MS cases and present in 25% and 17% of unaffected family members, respectively.

**Table 3.** Co-segregation of the 18 selected rare genetic variants with MS. Complete co-segregation is indicated as the variant being present in all MS cases and absent in all controls within one or two families, while acceptable co-segregation implies that the variant is present in all MS cases and is present in a maximum of 33% of controls. In total, 12 rare genetic variants show complete or acceptable co-segregation.

| Gene | Variant ID | Amino acid change | Co-segregation MS cases / controls (%) |
| --- | --- | --- | --- |
| *EXOC2* | rs760365995 | p.Pro134Leu | 100/50 |
| *PLK1* | rs35056440 | p.Leu261Phe | 100/0 |
| *MBP* | 18:74696828 | p.Gly151Val | 100/0 |
| *ZNF641* | rs200502528 | p.His342Tyr | 66/25 |
| *CCNI* | rs139547927 | p.Tyr334Cys | 100/66 |
| *ITPR3* | 6: 33648383 -<br>33648394 | - (DEL (a)) | 100/33 |
| *SORCS1* | 10:108439392 | p.Ser554Leu | 100/0 |
| *PRAM1* | rs138042924 | p.Ser387Pro | 100/20 |
| *CACNA1F* | X:49067910 | p.Met1324Leu | 100/29 |
| *MECP2* | rs61751445 | p.Thr311Met | 100/33 |
| *USH2A* | rs200802261 | p.Pro2870Leu | 75/17 |
| *CPT1A* | rs140958507 | p.Arg288Gln | 100/20 |
| *MTMR7* | rs760803217 | p.Thr6Met | 100/0 |

| Table 3 – continued. |  |  |  |
| --- | --- | --- | --- |
| Gene | Variant ID | Amino acid change | Co-segregation MS cases / controls (%) |
| *TRIM66* | rs138444298 | p.Arg1037Trp | 100/25 |
| *CNGB1* | rs776392588 | - (DEL (b)) | 100/50 |
| *TTC28* | 22:28501278 | p.Tyr1099Cys | 100/33 |
| *MANBA* | rs1283991082 | - (DEL (c)) | 100/50 |
| *TOX3* | 16:52580565 | p.Tyr24Cys | 100/0 |

Given the complex genetic etiology of MS, we did not expect a monogenic etiology in the families investigated here. Therefore, we argue that variants showing incomplete segregation are also of interest for further study. Investigating the function of the proteins encoded by the 12 genes that showed ‘acceptable’ co-segregation (presence in all MS cases and <33% of unaffected individuals) – *PLK1, MBP, ITPR3, SORCS1, PRAM1, CACNA1F, MECP2, CPT1A, MTMR7, TRIM66*, TTC28, and *TOX3* – we found that the proteins encoded by these genes could be linked to (de/re)myelination and/or the auto-immune response, as shown in the molecular framework in **Figure 1**.

**Figure 1.**
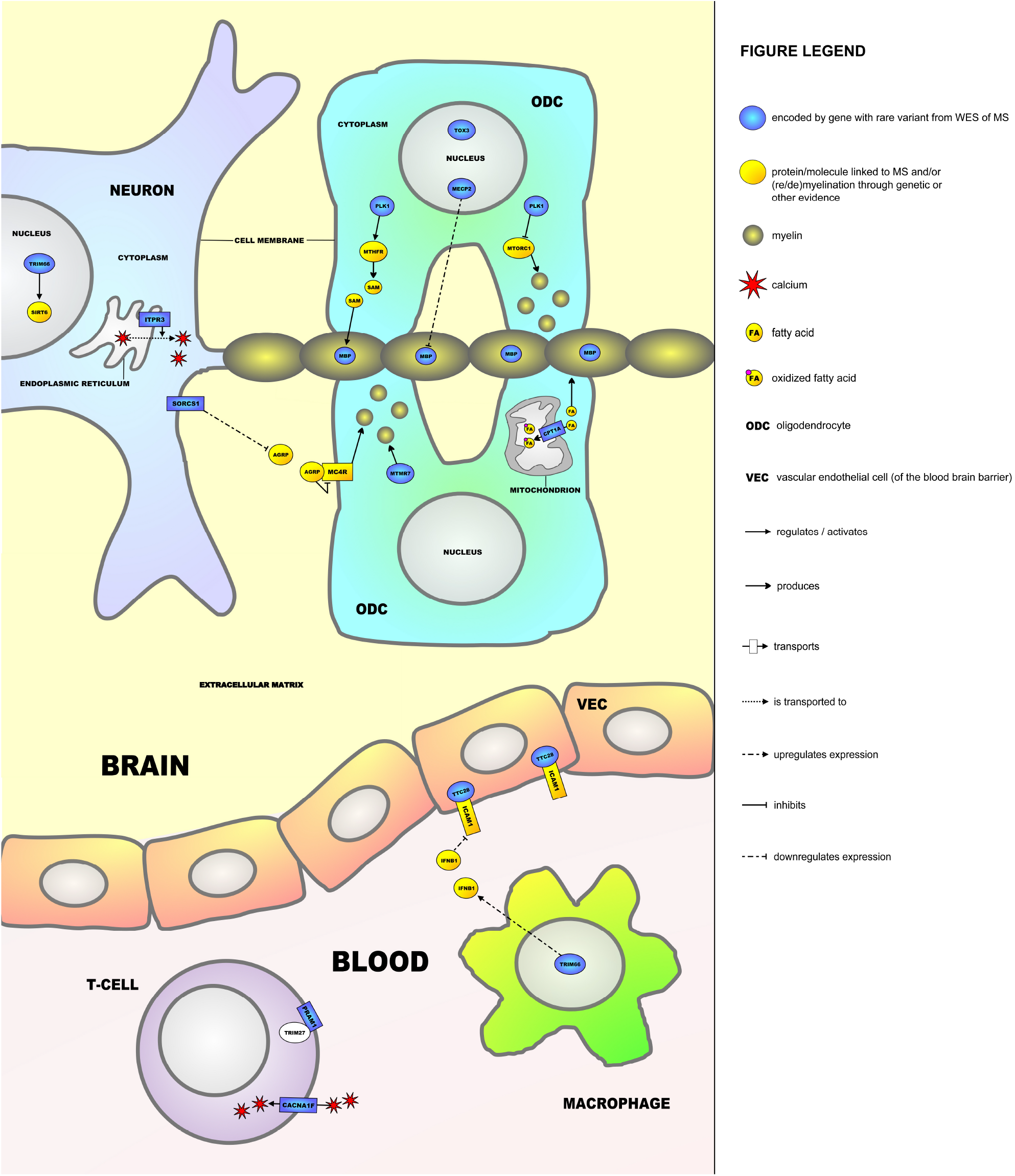
A molecular framework containing 12 interacting proteins encoded by genes harboring rare variants that were identified through whole exome sequencing (WES) in nine multi-incident multiple sclerosis (MS) families. MBP, PLK1, MECP2, MTMR7, TOX3, CPT1A, SORCS1, TRIM66, ITPR3, TTC28, CACNA1F, and PRAM1 are all linked to (de-/re)myelination and/or the auto-immune response characteristic of MS.

## Discussion

In the current study, we set out to identify rare genetic variants involved in MS through WES in multi-incident MS families. Among nine families, we identified rare variants in 12 genes that showed at least acceptable co-segregation with the disease. For each of these 12 genes, a link was identified with (de-/re)myelination processes and/or auto-immune responses that are characteristic of MS, and the encoded proteins could be placed in a molecular framework (**Figure 1**). Below, we will discuss how these 12 proteins – MBP, PLK1, MECP2, MTMR7, TOX3, CPT1A, SORCS1, TRIM66, ITPR3, TTC28, CACNA1F, and PRAM1 – and a number of additional molecules/proteins (most of which have also been linked to MS) operate in the framework.

MS is characterized by an auto-immune response against the myelin sheath that surrounds axons and is produced by the ODCs. Several genes in the molecular framework encode proteins that regulate myelination. Firstly, myelin basic protein (MBP) is the most important structural component of the myelin sheath. The most commonly used animal model of MS, experimental autoimmune encephalomyelitis (EAE), can be established through immunization (production of antibodies) of mice against Mbp ^47^. In this respect, antibodies against MBP have been found in EAE mice and a proportion of MS patients ^48^. In humans, anti-MBP antibodies were detected during the conversion of clinically isolated syndrome (CIS, a prodromal form of the disease) to clinical MS ^42^. Moreover, common genetic variation within *MBP* was found to predict the clinical course of MS ^43^.

Interestingly, in the most recently reported WES study of MS, a missense variant in *MBP* was found in four MS patients from the same family ^32^, further pointing towards a role of rare variants in *MBP* in MS susceptibility. The product of *PLK1* is a kinase that phosphorylates and regulates the activity of MTHFR, an enzyme is involved in the synthesis of methionine and its product S-adenosylmethionine (SAM) ^49,50^. SAM methylates MBP at arginine sites within the protein, which protects MBP against degradation and is hence important for maintaining the integrity of the myelin sheath ^51^. Common genetic variants in *MTHFR* have already been associated with the risk for MS development in multiple populations ^52-55^. PLK1 also inhibits the protein complex MTORC1, which in turn is an important positive regulator of (re)myelination by ODCs ^56,57^. The molecular framework gene *MECP2* encodes a transcriptional regulator that downregulates the expression of MBP in ODCs ^45,58^. Mecp2 has also been linked to negatively regulating (re)myelination in the EAE mouse model ^44,59^. *MTMR7* encodes a member of the myotubularin protein family of phosphatases and is upregulated after ODC differentiation, which may point to a role in (re)myelination ^60^. *TOX3* encodes a transcription factor that positively regulates the survival of ODCs after injury and may therefore be involved in remyelination ^61^. Another important protein in the framework is the product of the *CPT1A* gene, an enzyme located in the mitochondrial membrane that catalyzes the rate-limiting step in fatty acid oxidation. When oxidized, fatty acids cause oxidative stress and cannot be used for myelin production, which has a negative effect on (re)myelination by ODCs ^46,62^. In two isolated, indigenous populations from northern Canada - the Inuit and Hutterites - two different rare genetic variants in *CPT1A* occur in the majority of the population. As people carrying either one of these variants have a much smaller chance to develop MS compared to the nonindigenous population of Canada, these variants seem to protect against the disease. The two variants result in 78% decreased activity and complete inactivity of CPT1A in the Inuit and Hutterite populations, respectively ^46^. This study also showed that in a mouse model of the low-activity Inuit-variant in *CPT1A*, EAE could not be induced, further providing evidence for a protective role for this variant.

The molecular framework gene *SORCS1* encodes a membrane protein that is highly expressed in neurons and is involved in downregulating the expression of agouti-related peptide (AGRP) ^63,64^. AGRP has been linked to MS and is a secreted molecule that binds to and is an antagonist of the melanocortin-4 receptor (MC4R) ^65-67^. MC4Rs are highly expressed in ODCs and their precursor cells. When bound by their main agonist, the melanocortin peptide ACTH (not shown in **Figure 1**), MC4Rs have been shown to protect ODCs from damaging insults and increase proliferation of their precursor cells, hence contributing to (re)myelination ^68^. TRIM66 in the framework is a transcription factor that is highly expressed in both neurons and macrophages. Interestingly, two rare variants in *TRIM66* have been associated with MS severity ^69^. In neurons, TRIM66 regulates the activity of SIRT6, a histone-deacetylating enzyme that is involved in epigenetic processes and has been recently suggested as a novel drug target for MS ^70,71^. In (peripheral) macrophages, TRIM66 is involved in upregulating the expression of endogenous interferon beta (IFNB1) ^72^, which is interesting as recombinant, exogenous IFNB1 is one of the earliest and still used disease-modifying treatments for MS ^73^. Lastly, ITPR3 is a protein in the endoplasmic reticulum (ER) membrane that increases the release of calcium from the ER to the cytoplasm of neurons. This triggers a downstream signaling cascade that can eventually result in neuronal death/neurodegeneration in both the EAE mouse model of MS ^74^ and stem cell-derived neurons from MS patients ^75^, as sustained high calcium levels in cells/neurons are highly toxic.

In addition to TRIM66 - which functions both in neurons and peripheral macrophages - three proteins encoded by genes with MS-related rare variants found in our study operate in the framework outside of the CNS. First, TTC28 is a cytoplasmic protein located in vascular endothelial cells that constitute the blood-brain barrier (BBB) between the periphery and the CNS. TTC28 binds and functionally interacts with ICAM1 ^76^, a membrane receptor that has been consistently linked to the pathogenesis of MS, as it promotes transcellular diapedesis of inflammatory T-cells from the periphery to the brain cells across the BBB, a key pathological process underlying MS ^77^. Moreover, exogenous IFNB1 administration leads to downregulation of ICAM1 in vascular endothelial cells of the BBB ^78^.

The two other identified genes encode proteins with functions in peripheral T-cells. *CACNA1F* encodes an L-type voltage dependent calcium channel that transports calcium into T-cells. A deficiency of CACNA1F leads to reduced calcium influx and ultimately disturbances in T-cell function that are characteristic of MS ^79-81^. PRAM1 is a membrane protein that binds and interacts with TRIM27, a cytoplasmic protein that negatively regulates calcium influx into T-cells, with a similar effect on T-cell function as CACNA1F deficiency ^82,83^.

In summary, 9 of the 12 genes with rare variants associated with MS in our study encode proteins that are (potentially) involved in regulating (de-/re-)myelination (*MBP, PLK1, MECP2, MTMR7, TOX3, CPT1A, SORCS1, TRIM66*, and *ITPR3*), while 4 of the 12 genes (*TRIM66, TTC28, CACNA1F*, and *PRAM1*) are linked to (auto-)immune processes. Although they need to be replicated/validated (see below), our findings may imply that rare variants predominantly affect genes involved in (de-/re-)myelination and, eventually, neurodegeneration rather than the acute auto-immune reaction that is associated with disease relapses.

Our study should be viewed in the context of a number of strengths and limitations. A strength is the fact that many of the genes found affected are supported by evidence from previous studies of rare and common genetic variants. One important limitation is the small number of family members for which co-segregation analyses of the identified variants could be performed. Larger families and a larger numbers of participants would yield power for statistical analysis of these co-segregation analyses. Future research should therefore be aimed at including more family members in this type of studies. Moreover, 7 of the 12 identified variants did not show complete co-segregation with MS. For instance, unaffected subject I:2 in family 1 (**Supplementary Figure 1**) carries the identified missense variant in *EXOC2*, which led us to not further consider this variant for the framework, as it is found in 1 out of 2 tested unaffected family members. However, no variants in *PLK1* and *MBP* are found in subject I:2 that are present – together with the *EXOC2* variant - in all three MS patients from this family: II:5, II:11 and II:13.

Further, none of the three variants were found in the second unaffected member of the family, II:4. Another example is the identified missense variant in *CACNA1F* that was found in families 3 and 7 (**Supplementary Figure 1**); in both these families, one unaffected family member also carried this variant (i.e. III:3 and II:9, respectively). This means that, across both families, 2 out of 7 unaffected family members (29%) carry the *CACNA1F* variant. However, individual III:3 from family 3 may still be at risk of developing MS, as she is only in her 30s. These patterns of incomplete co-segregation suggest an oligogenic inheritance of MS with the effects of a small(er) number of variants still needed for the MS phenotype to be fully expressed.

As indicated above, a number of genes that were identified in this study have been previously linked to MS, and it would be of interest to perform functional experiments involving these or other identified genes to further elucidate the (pathogenic) molecular mechanisms underlying MS. For instance, in the study by Mørkholt et al., it was shown that a rare variant in *Cpt1a* results in ineffective EAE inducement in mice ^46^. In this respect, it could be speculated that the *CPT1A* variant that was identified in this study leads to a gain-of-function of the CPT1A enzyme, and it could be tested whether this is indeed the case through *in vitro* studies.

In conclusion, through WES and subsequent Sanger sequencing in multi-incident MS families, rare genetic variants in 12 genes were identified that show an acceptable co-segregation pattern with the disease. The proteins encoded by these genes interact in a molecular framework that contains both (de/re)myelination and auto-immunity-related processes. Our findings provide further insights into the molecular mechanisms underlying MS that should be studied in cellular and/or animal models, and that could be leveraged for the development of novel treatment strategies for the disease, and this specifically aimed at promoting remyelination.

## Supporting information

Horjus et al. Supplementary Table 1

## Data Availability

All data produced in the present work are contained in the manuscript

## Acknowledgements

We would like to thank all the MS patients and their unaffected family members who have contributed to this study. In addition, we are grateful to professor Christian Gilissen and his research group at the Department of Human Genetics, Radboud University Medical Center, Nijmegen, The Netherlands, for their assistance with the analysis and interpretation of the WES data.

---

**Supplementary Table 2.**
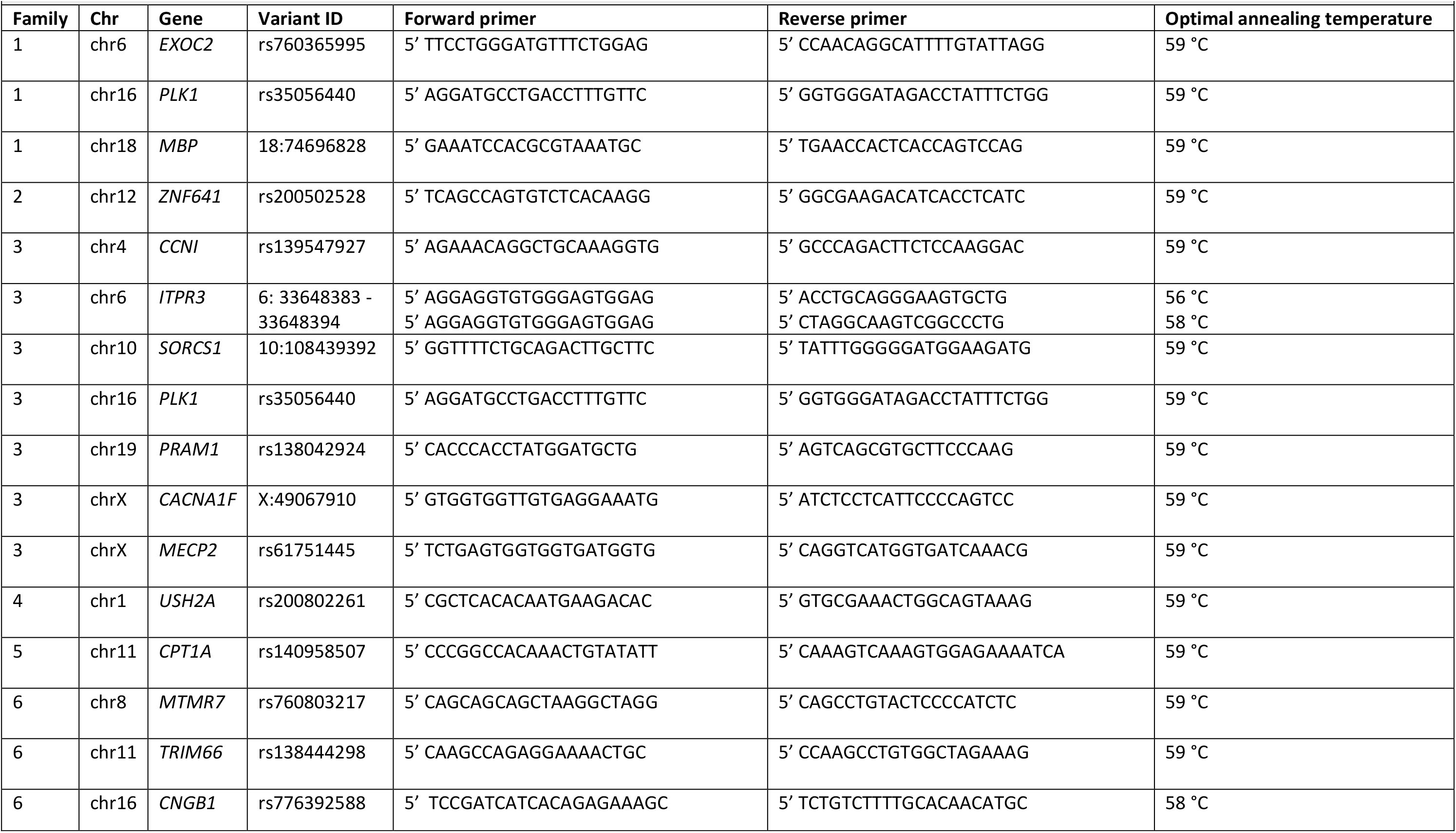
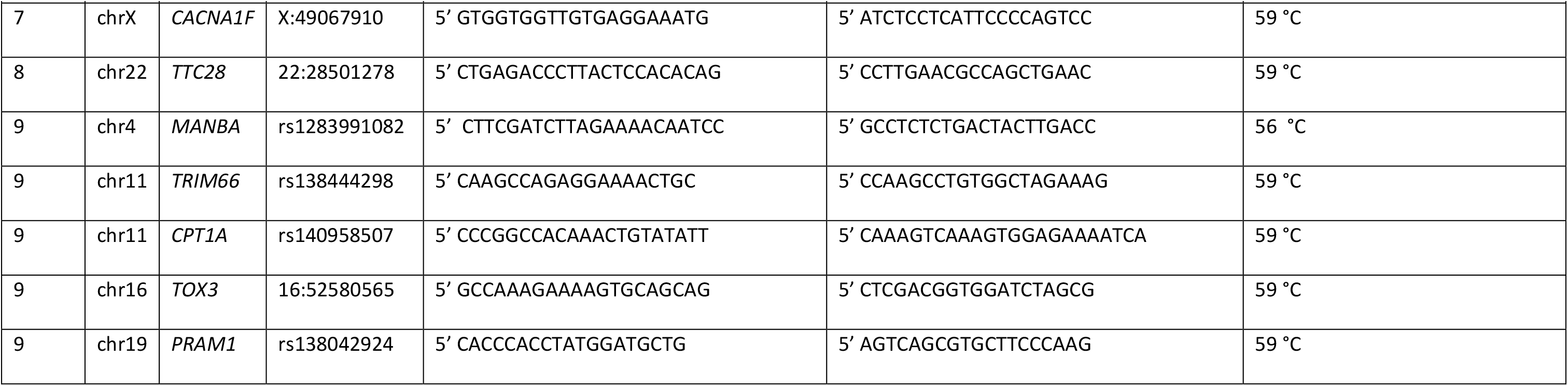
An overview of the sequences of the forward and reverse primers that were used for the Sanger sequencing and their optimal annealing temperatures in order to validate the 18 variants in the family members of the multi-incident MS families.

**Supplementary Table 3.** An overview of the selection criteria that each of the 18 rare genetic variants meets is provided. If a variant or gene had a significant P-value in either the exome chip study ^24^ or based on the results from the genome-wide association study (GWAS) ^23^ – i.e. the gene-wide P-value – by the International Multiple Sclerosis Genetics Consortium (IMSGC), said P-value is provided.

| Gene | Variant ID | Amino acid change | Present in 2/9 families | Literature link(s) with MS | P-value exome chip | Gene-wide P-value |
| --- | --- | --- | --- | --- | --- | --- |
| *EXOC2* | rs760365995 | p.Pro134Leu |  |  |  | 2.50E-03 |
| *PLK1* | rs35056440 | p.Leu261Phe | ✓ |  |  |  |
| *MBP* | 18:74696828 | p.Gly151Val |  | ✓<sup>42,43</sup> |  |  |
| *ZNF641* | rs200502528 | p.His342Tyr |  |  |  | 3.76E-02 |
| *CCNI* | rs139547927 | p.Tyr334Cys |  |  |  | 1.46E-03 |
| *ITPR3* | 6: 33648383 - 33648394 | - (DEL (a)) |  |  |  | 2.65E-05 |
| *SORCS1* | 10:108439392 | p.Ser554Leu |  |  |  | 3.74E-02 |
| *PRAM1* | rs138042924 | p.Ser387Pro | ✓ |  | 3.26E-02 | 3.55E-04 |
| *CACNA1F* | X:49067910 | p.Met1324Leu | ✓ |  |  |  |
| *MECP2* | rs61751445 | p.Thr311Met |  | ✓<sup>44,45</sup> |  |  |
| *USH2A* | rs200802261 | p.Pro2870Leu |  |  | 2.76E-02 |  |
| *CPT1A* | rs140958507 | p.Arg288Gln | ✓ | ✓<sup>46</sup> |  |  |

| Supplementary Table 3 – continued. |  |  |  |  |  |  |
| --- | --- | --- | --- | --- | --- | --- |
| Gene | Variant ID | Amino acid change | Present in 2/9 families | Literature link(s) with MS | P-value exome chip | Gene-wide P-value |
| *MTMR7* | rs760803217 | p.Thr6Met |  |  |  | 2.83E-02 |
| *TRIM66* | rs138444298 | p.Arg1037Trp | ✓ |  |  |  |
| *CNGB1* | rs776392588 | - (DEL (b)) |  |  |  |  |
| *TTC28* | 22:28501278 | p.Tyr1099Cys |  |  |  | 3.15E-07 |
| *MANBA* | rs1283991082 | - (DEL (c)) |  |  |  |  |
| *TOX3* | 16:52580565 | p.Tyr24Cys |  |  |  | 3.07E-02 |

